# The relationship between health literacy and adherence to physical activity guidelines: a scoping review

**DOI:** 10.1101/2024.10.22.24315903

**Authors:** Alex Lawrence, Jon Wardle, Jacqui Susan Yoxall

## Abstract

**Objective:** To examine the relationship between health literacy and adherence to physical activity guidelines.

**Methods:** In line with the Joanna Briggs Institute framework, we conducted a scoping review of the literature.

**Results:** Out of 2,098 articles identified, 19 met the inclusion criteria. The studies utilised various health literacy measurement tools, with the European Health Literacy Survey being used the most. Fifteen studies examined the association between total health literacy scores and achieving >150 minutes of moderate-to-vigorous physical activity weekly. Nine studies reported a positive association, while others found no significant association. In particular, studies using self-reported physical activity more frequently found an association, whereas no association was found when using objective physical activity measures.

**Conclusions:** The findings of this review were inconclusive. The lack of standard health literacy instruments and reporting presents a barrier to the field of knowledge progressing. Moreover, longitudinal relationships between health literacy, mediators and physical activity need to be investigated.

## Introduction

Health literacy (HL) refers to an individual’s capacity to navigate, understand, and use health-related information to manage their health.^1,2^ Low health literacy is linked to poor health outcomes, including higher mortality rates, medication non-adherence, and difficulties navigating health systems,^3–6^ and increased risk of unhealthy behaviours, such as physical inactivity.^5^ Such associations make HL particularly interesting for public health and health promotion efforts.

Approximately one-third (31%) of adults, or 1.8 billion people, are physically inactive.^6^ Physical inactivity is a major modifiable risk factor of all-cause mortality, accounting for 6% of premature deaths worldwide.^7,8^ Increasing physical activity (PA) prevents, mitigates, and ameliorates the effects of many non-communicable diseases, injuries, and associated comorbidities.^9^ Furthermore, achieving the recommended levels of PA can reduce the risk of all-cause mortality by 75%.^10^ While meeting the PA guidelines is attainable for most individuals, adopting and maintaining an active lifestyle also requires significant cognitive skills to overcome motivational and scheduling barriers.^11^ As efforts to achieve global PA targets continue, the nexus between HL and adherence to PA guidelines is becoming increasingly relevant.

Previous systematic reviews have concluded that there is a positive relationship between HL and PA levels across different age groups.^12,13^ However, these findings referred to increase in PA irrespective of whether such an increase meets recommended PA guidelines. Although any increase in PA is broadly beneficial, public health policy and surveillance are primarily concerned with adherence to PA guidelines. By focusing on PA guidelines, the outcomes of this study align with public health priorities, targeting the relationship between HL and the threshold of PA most likely to result in meaningful health improvements.

The variation in PA and HL measurement tools, and the complex nature of the HL construct generally^14, 15^ make comparing and synthesising findings across studies challenging. Scoping reviews, however, are particularly valuable in such cases, as they are well-suited for literature that is heterogeneous.^16^ Scoping reviews aim to provide an overview of an area or issue.^17^ They can help identify knowledge gaps, evaluate the breadth of literature, develop areas of inquiry, or review prior research methods.^18^ For example, scoping reviews can inform systematic reviews about whether the inclusion criteria or research questions are appropriate. A scoping review of the relationships between PA and HL would allow for a broader review of quantitative, qualitative, and mixed-methods research across various academic disciplines (E.g., public health, education, psychology, and sports science) to highlight areas lacking research. To our knowledge, no scoping review on HL and PA has been published.

This scoping review aims to identify and synthesise research on the relationships between HL and adherence to PA recommendations. Specifically, the proposed review seeks to answer the following review questions: (i) What is the association between HL and adhering to PA guidelines in the general adult population?; (ii) What HL constructs are being measured, and what instruments are used to assess them?; (iii) How does implementing HL interventions compare to no intervention or standard care impact adherence to PA guidelines in the general adult population?; and (iv) What are the patient and social-level factors suggested to influence the connection between HL and health numeracy, and adhering to PA guidelines?

## Materials and Methods

This scoping review followed the Joana Briggs Institute (JBI) approach for scoping reviews.^18^ The JBI approach was used for its methodological quality and the availability of tools to guide scoping reviews.^19^ The research team developed the review protocol and registered with the Open Science Framework on the 20^th^ of February 2024 (https://osf.io/smqfj). This report was written using the Preferred Reporting Items for Systematic Reviews and Meta-Analyses Extension for Scoping Reviews (PRISMA-ScR) checklist.^20^

To identify the key concepts relevant to the primary research question, “What is the association between health literacy and physical activity in the general adult population?” we used the Population, Concept, and Context (PCC) framework recommended by the Joanna Briggs Institute.

### Eligibility criteria

We included observational or intervention studies written in English and involved the general adult population (18 years or older). The restriction to studies written in English was due to resource constraints. Studies with adults and children were considered if they reported data separately for adults. However, studies focused exclusively on adolescents or children were excluded.

Eligible research studies had to use a validated measure of HL. Self-assessed (i.e., subjective) measures, where participants estimate their HL, were excluded as these assessments are generally intended for use in clinical settings rather than research purposes. For intervention studies, HL needed to be measured both at baseline and after the intervention and demonstrate improvements in the post intervention measures.^21^

To qualify for inclusion, PA needed to be reported as either a primary or secondary outcome, and groups need to be dichotomised as physically inactive and PA or similar. For this study, people who do not meet recommended levels of regular PA were defined as “physically inactive”. Where pedometers were used to measure PA, participants must achieve at least 7,000 steps per day to be considered sufficiently active.

### Information sources and search

The search strategy followed the Peer Review of Electronic Search Strategies standard.^22^ An initial search on PubMed was performed to identify relevant articles based on keywords in titles, abstracts, and index terms. Subsequently, a Health Sciences Librarian assessed the strategy to balance breadth, comprehensiveness, and feasibility. A search strategy was then tested on PubMed to verify and refine the search query as needed to ensure it captured a significant number of relevant studies. Once finalised, the strategy was adjusted to suit other databases’ syntax and subject headings. Validation and updates will continue throughout the review process to maintain the effectiveness of the PubMed search strategy. The following bibliographic databases were searched: MEDLINE, ProQuest, Scopus, CINAH, Web of Science (Core Collection), PubMed, and PsycINFO. The search was unrestricted by date. To achieve literature saturation, we will examine the reference lists of included studies and relevant reviews identified during the search process.

### Information sources and search

The results of each review were exported to EndNote 20 (Clarivate Analytics, PA, USA) to remove duplicates. The remaining articles were imported into Covidence (www.covidence.org) for further duplicate detection. One reviewer screened the titles and abstracts; potentially relevant articles were retrieved in full and added to Covidence. Two independent reviewers assessed the full texts against the inclusion criteria, with any exclusions documented according to JBI guidelines. Discrepancies were resolved through discussion or by consulting an additional reviewer. The search results and study inclusion process will be detailed in the final scoping review and presented using a PRISMA-ScR flow diagram.^20^

### Data Extraction

The Joanna Briggs Institute standardised data extraction form guided data extraction from each included study. The data extraction form was piloted on a small subset of studies to ensure clarity, completeness, and consistency in data collection. The form was refined based on issues identified during this pilot phase. Data extraction was performed independently by one reviewer (AL). The reviewers were not required to contact study authors or investigators to clarify any ambiguous or missing data. The reason for exclusion has been noted and described in the PRISMA flow diagram (Figure 1). Consistent with guidance on scoping reviews, we did not perform any methodological quality appraisal of the included reports.

**Figure 1.**
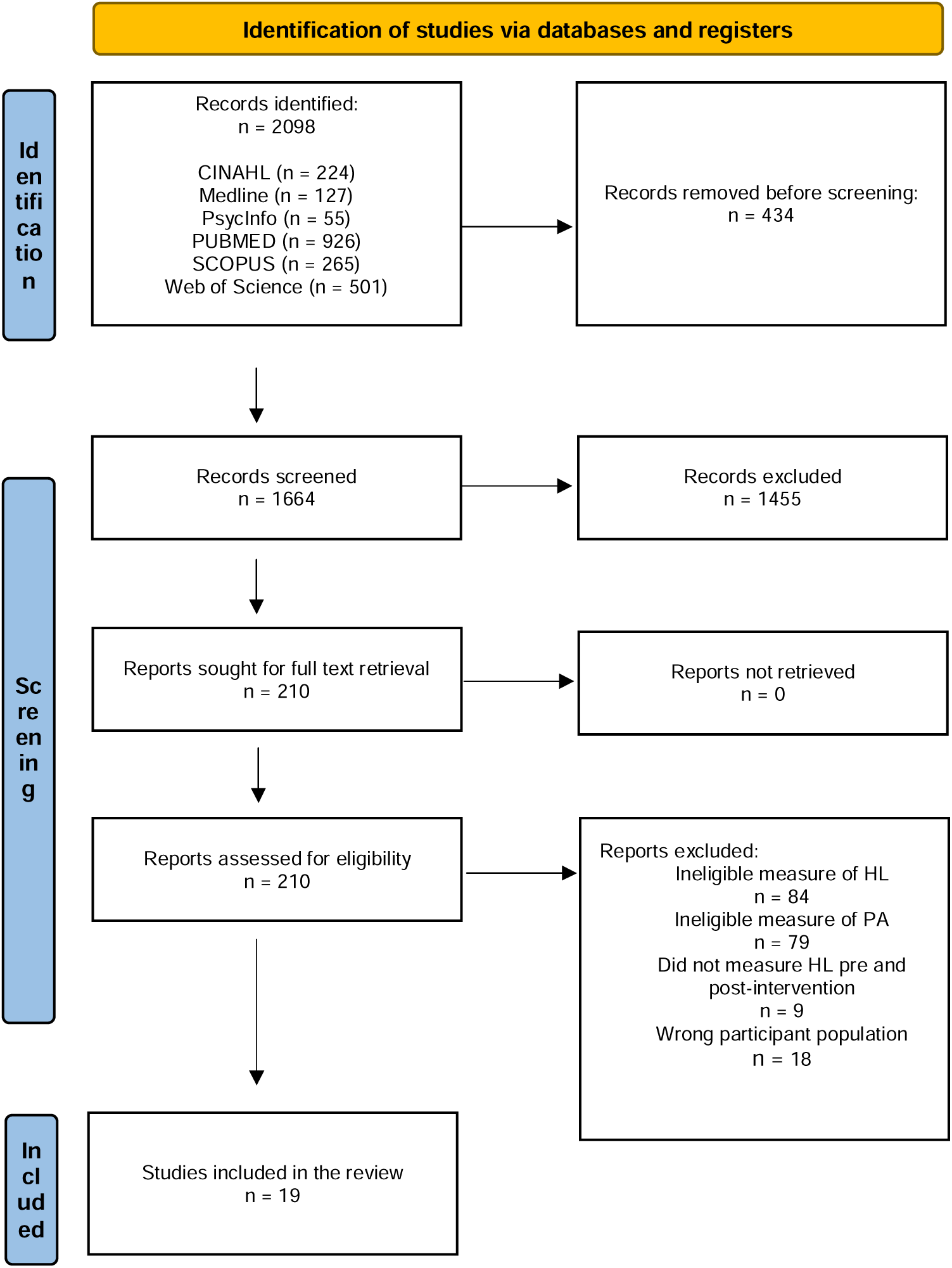
Flow diagram of article selection process.

### Data analysis

A numerical analysis of the nature and extent of the studies was performed. A narrative summary supplemented the analysis to synthesise the main themes, patterns, and conclusions from the review, explaining how the results align with the review’s aim and questions.

## Results

The selection of sources of evidence is illustrated in Figure 1. A total of 2,098 articles were identified from a search of electronic databases. After removing 434 duplicates, 1,664 titles and abstracts were screened. Full texts were retrieved for 210 studies, of which 191 were excluded for reasons such as not adequately measuring HL (n = 84), did not measure HL pre and post-intervention (n = 10), including the wrong participant population (n = 18), and issues with validity, alignment with guidelines, and classification of PA (n = 79). A total of 19 articles met the inclusion criteria and were included in the review.

### Study characteristics

The study characteristics are summarised in Table 1. Among the studies selected for inclusion, most were cross-sectional design (n = 13),^23–35^ followed by longitudinal design(n=3),^36–38^ prospective cohort design (n = 1),^39^ correlational study (n = 1),^40^ a combined cohort and case-control methods (n = 1).^41^ Most studies were published in Europe (n = 10),^26,28,30,31,35–39,41^ including Germany (n = 2),^30,37^ Denmark (n = 2),^31,39^ Netherlands (n = 2),^36,41^ United Kingdom (n = 1),^38^ Italy (n =1),^35^ Ireland (n = 1),^26^ and a multi-national European study (n = 1).^28^ Otherwise, studies were published in Canada (n = 1),^23^ Australia (n = 2)^27,29^ Turkey (n = 2),^25,40^ and there was one study each from Iran,^34^ Japan,^32^ Singapore,^24^ and the United States of America.^33^

INSERT TABLE 1

### Health literacy

Various HL assessment tools were used in the included studies. Of the 19 studies, 3 used only a portion of a survey,^28,38,39^ Most studies used a version of the European health literacy survey (HLS-EU), including the HLS-EU-Q16 (n = 4),^25,30,31,37^ HLS-EU-Q47 (n = 1),^26^ and the Italian version of the HLS-EU-Q6 (n = 1),^35^ The Brief Health Literacy Screen was utilised in 4 studies,^23,24,36,41^ Two studies used the Health Literacy Questionnaire (HLQ-9); however, neither study used the entire scale. Specifically, Brors et al.^39^ used 4 of the 9 measurements: accessing, understanding, and appraising health information and the availability of social support to assist in managing health. Jennings et al.^28^ only measured understanding of health information. The Health Literacy Management Scale^27,29^ was used in 2 studies. The Turkish Health Literacy Scale-32,^40^ the Newest Vital Sign,^40^ the Organisation for Economic Co-operation and Development International Adult Literacy Survey,^38^ the Health Literacy for Iranian Adults tool,^34^ the Medical Term Recognition Test,^33^ the Functional, Communicative, and Critical Health Literacy Scale^32^ featured in one study each.

### Physical activity

Eighteen studies relied on self-reported measurements of PA. Of these, 12 used a direct question,^25–30,35,37–41^ 1 used the Global Physical Activity Questionnaire,^24^ 1 used the SQUASH questionnaire,^36^ 1 used the SDSCA questionnaire,^31^ and 2 employed the IPAQ short form.^32,34^ Additionally, one study utilised the Health Information National Trends Survey.^33^ One study combined both an objective measure, using a pedometer, with a self-reported measure of PA, specifically the Leisure Score Index from the Godin Leisure-Time Exercise Questionnaire.^23^

### Findings

Fifteen studies examined the association between *total* HL scores and PA of ≥ 150 minutes of MVPA per week. Nine studies found a significant positive association between PA and HL, ^23,25,27–29,34–36,41^ while 4 studies found no such association.^24,30,31,33^ One study reported a significant association using unadjusted odds ratios, but this association became non-significant after adjusting for covariates.^38^ Another study found that individuals with *inadequate* HL were more likely to engage in sufficient PA.^40^

One study specifically explored the relationship between PA of ≥ 600 MET and HL.^24^ The unadjusted model indicated a significant association, but this association was no longer significant after adjustment for covariates. No association was found between adequate HL and the likelihood of achieving sufficient PA (≥ 8000 steps/day) in either crude or adjusted models.

Five studies investigated the relationship between PA of ≥ 150 minutes per week and a specific subdomain of HL. One study consistently found significant associations between PA and the subdomains of social support, appraisal, access, and understanding in both crude and adjusted models.^34^ Communicative HL was found to have a *non-significant* association with PA in 2 studies using unadjusted models; however, both studies reported no association when using adjusted models,^31,32^ Three studies examined functional HL. Two reported non-significant associations in crude models,^31,32^ and one found no association.^33^ Only two of these studies used adjusted models, and both reported no association.^31,32^ One study focused on three HL domains: Disease Prevention, Health Promotion, and Healthcare, each with four sub-indices: Accessing, Understanding, Evaluating, and Applying. Significant findings were reported only for Health Promotion (combined), Disease Prevention: Understanding, Health Promotion: Accessing, Health Promotion: Understanding, and Health Promotion: Evaluating.^26^

### Pathways

One study investigated if self-efficacy, neutral/negative attitude and low-risk perception mediated the relationship between HL and sufficient PA.^36^ Self-efficacy mediated 32% of the association between HL and sufficient PA, while neutral/negative attitude and low-risk perception did not show as mediators.^36^

## Discussion

### Summary of Key Finding

This scoping review aimed to identify and synthesise existing research on the relationship between HL and PA This review builds on previous work by Buja ^12^ and Lim et al. ^13^ by explicitly focusing on HL and its relationship with adherence to PA recommendations. The findings of this systematic review were inconsistent with those of Buja ^12^ and Lim et al. ^13^ This discrepancy may be in part caused by previous studies overlooking important research. Several studies not documented by Buja^12^ were identified in this review, including those conducted by Gibney and Doyle,^26^ Jayasinghe et al.^27^ Jennings et al,^28^ Juul et al,^31^ and Reider et al.^33^ Considering that our search, which has a narrow focus on PA guidelines, identified missed studies, it is plausible that previous evaluations may have overlooked further research outside our review’s scope. Furthermore, half of the studies included in this review were published after Buja.^12^ Considering the concept of HL is changing and increasing in complexity, more recent studies might be indicating a shift in the relationship between HL and PA. Therefore, this review may provide a new viewpoint on the impact of HL on PA, reflecting the changing nature of HL itself. Nevertheless, the changing concept of HL is improbable in explaining the disparity in our results, as Buja^12^ and Lim et al^13^ also documented substantial variation in the measure of HL between studies.

The most likely explanation for the variation in results is in the measure of the PA. This study specifically reviewed adherence to PA guidelines, while other studies included a broader range of PA measures, including PA level (low, moderate, high), mean PA, and PA intensity (light, moderate, vigorous). PA guidelines are designed to be understood and followed, reducing an individual need to make decision-making compared to more self-directed forms of PA. As such, the complexity of PA tasks, in this case, the simplicity of following PA guidelines, are likely key factors in explaining the differences in findings.

The results of our review suggest that there is some evidence to support an association between HL and PA. Compared with individuals with low HL, people with adequate HL were more likely to comply with PA recommendations and maintain a more active lifestyle. Nevertheless, all of these studies depend on self-reported PA data, which is susceptible to recall bias and inaccuracies in documenting actual PA.^42^ This implies the necessity of exercising caution when interpreting the results and restricts the reliability of the findings. Therefore, although some evidence suggests an association between HL and PA, the dependence on self-reported data introduces constraints that must be resolved in subsequent research.

Interestingly, the only study that used an objective measure of PA reported no association between HL and PA.^23^ This is a key finding that contrasts with studies relying on self-reported data. An explanation for this may be the role of self-perception in HL. Individuals with higher HL might report PA levels that align more with health recommendations, even if their actual activity levels do not differ significantly from individuals with lower HL when measured objectively. Another explanation is HL may influence how PA is reported in self-report questionnaires. Participants with inadequate HL might have underreported their actual PA compared to those with adequate HL, potentially due to misunderstanding or misinterpreting the questions. Overall, the evidence on the relationship between HL and PA remains inconclusive. While there is some indication of a link, the current body of research is insufficient.

### Gaps in the literature and future recommendations

Most studies are cross-sectional, limiting conclusions about causality between HL and PA. This limitation arises because cross-sectional designs capture data at a single point, making it difficult to track changes and effects over time.^43^ Longitudinal or intervention studies would providing stronger evidence for causation. Future studies should prioritise longitudinal studies to progress knowledge of the interaction between HL and adherence to PA guidelines. Moreover, none of the intervention studies satisfied the inclusion requirements, indicating a gap in the research that should be addressed.

Half of the studies in our review used multivariate logistic regression analysis. This approach enhanced the clarity of the results by accounting for various confounders and allowed for the measurement of odds ratios, which quantifies the strength of the association between each predictor and the outcome, making the results easier to interpret.^44^ However, the selection of confounding variables varied across studies, which complicated synthesising the overall findings. Furthermore, some studies controlled for factors that may lie within the causal pathway. Factors like knowledge and income might mediate the relationship between HL and health outcomes. This “overadjustment” could have underestimated the true effect of HL by masking its direct influence on PA.

Only one study in this review investigated possible mediating effects, identifying self-efficacy as mediating the relationship between HL and PA.^36^ This finding is consistent with previous empirical studies that found that self-efficacy mediates HL and self-care behaviours.^45^

### Implications

The findings from this review reinforce the need for further research to better understand the association between HL and PA. The review’s inconclusive results suggest that there is not yet a consistent link between HL and compliance with PA guidelines. This raises concerns about assuming that improving HL alone will significantly enhance PA. Policymakers should therefore be cautious when relying on HL as the primary driver of interventions aimed at raising population PA levels to meet guidelines. In practice, healthcare professionals should continue to acknowledge HL in the provision of services. However, when implementing strategies to improve PA adherence, focusing on approaches with stronger evidence-based behavioural outcomes will offer a more comprehensive solution for enhancing PA compliance.

### Strengths and Limitations

Our study has several strengths. First, the extensive search strategy makes it unlikely that any significant studies were missed. Additionally, the low risk of bias from selective reporting strengthens the study’s objectivity, as many published studies show no relationship between HL and compliance with PA recommendations. However, there are some limitations. This review did not assess the methodological quality of the selected studies restricts our ability to assess the overall findings. Furthermore, including only English-language articles may have excluded important studies published in other languages, potentially introducing a language bias and limiting the generalisability of the results.

## Conclusion

To our knowledge, this is the first scoping review exploring the association between HL and PA. Despite the growing prevalence and importance of HL research, Future research should focus on longitudinal designs, standardising HL measures, and exploring the potential mediators of the HL-PA relationship, such as self-efficacy.

## Data Availability

All data produced in the present work are contained in the manuscript

## Acknowledgements

Declaration of Interest Statement: The authors reported No potential competing interest. Funder Information: No funding was obtained for this project

## Notes

### Competing Interest Statement

The authors have declared no competing interest.

### Clinical Protocols

https://osf.io/a8uw6

### Funding Statement

This study did not receive any funding

### Summary of Updates

The original manuscript contained an intervention study that did not meet the study's eligibility criteria.

