## Supplementary material for "The relationship between health literacy and adherence to physical activity guidelines: a scoping review": Table 1

**Summary of study designs, participants, and intervention procedures**

| **No** | **Authors,**  **Year of Publication,**  **Country** | **Research question** | **Study design**  **Population** | **Health literacy Scale**  **Health literacy**  **domains being measured** | **Measure of PA** | **Results** |
| --- | --- | --- | --- | --- | --- | --- |
| 1 | Ahcioglu and Yilmazel  2022  Turkey | To examine the health literacy and associated factors among coronary artery disease patients. | Correlational study  275 Hospitalised patients (≥ 50 years) diagnosed as any CVD in the cardiology service and coronary intensive care unit. | Turkish Health Literacy Scale-32 (TSOY-32)  Not mentioned | Self-reported measure of PA with data being gathered via face-to-face interviews. | 51.5% of participants with inadequate HL were sufficiently PA (≥ 30 min/d), compared to 48% participants with inadequate HL who were insufficiently PA (< 30 min/d). The results were significant (χ^2^=7.5000, P<0.05). |
| 2 | Al Sayah et al.  2016  Canada | To investigate whether inadequate health literacy is associated with self-reported moderate to vigorous physical activity in community-dwelling adults. | Cross sectional study  1296 Participants | Three validated questions.  (1) Problems learning;  (2) Confidence filling out medical forms;  (3) Help reading. | Self-reported measure of PA using the Lesure Score Index of the Goldin Leasure-Time Exercise Questionnaire to estimate physical activity  Objective measure using pedometer. | Crude model:  A significant association was found between participants with adequate HL and likelihood to achieve sufficient PA as measured by ≥ 150 mins/wk (OR 0.51, 95% CI 0.34-0.77, p = 0.001); and ≥ 600 MET/wk (OR 0.53, 95% CI 0.34-0.80, p = 0.003).  No association was found between participants with adequate HL and likelihood to achieve sufficient PA (≥ 8000 steps/d) (OR 1.02, 95% CI 0.59-0.1.77, p = 0.94)  Adjusted model*:  A significant association was found between likelihood to achieve sufficient PA (≥ 150 mins/wk) and adequate HL (OR_adj_ 0.63, 95% CI 0.41-0.97, p = 0.037).  A non-significant association was found between likelihood to achieve sufficient PA (≥ 600 MET/wk) and adequate HL (OR_adj_ 0.65, 95% CI 0.42-1.01, p = 0.06).  No association was found between likelihood to achieve sufficient PA (≥ 8000 steps/d) and adequate HL (OR_adj_ 1.27, 95% CI 0.70-2.29, p = 0.43)  *Adjusted for age, gender, education, income, employment, ethnicity, smoking status, BMI, and number of chronic illnesses. |

Table 1. (continued)

| **No** | **Authors,**  **Year of Publication,**  **Country** | **Research question** | **Study design**  **Population** | **Health literacy Scale**  **Health literacy**  **domains being measured** | **Measure of PA** | **Results** |
| --- | --- | --- | --- | --- | --- | --- |
| 3 | Asharani et al.  2021  Singapore | To examine the functional health literacy of the nation, factors associated with health literacy, and its relationship with diabetes recognition. | Cross sectional study  2895 participants | The Brief Health literacy Screen  (1) Problems learning;  (2) Confidence filling out medical forms;  (3) Help reading. | Self-reported measure of PA using the Global Physical Activity Questionnaire (GPAQ) | 84.4 % of participants with adequate HL were sufficiently PA (≥ 150 mins/wk), compared to 79.3% of participants with inadequate HL who were insufficiently PA (< 150 mins/wk). The results were not significant (P = 0.06).  Adjusted model*:  No association was found between likelihood to achieve sufficient PA (≥ 150 mins/wk) and adequate HL (OR 1.1, 95% CI 0.7-1.6, p = 0.78)  *Sociodemographic variables |
| 4 | Brors et al.  2022  Denmark | To investigate the associate between health literacy, behavioural and psychologic risk factors, BMI, physical activity, anxiety depression, and smoking status in patients treated with percutaneous coronary intervention, as well as to assess changes in these factors over a 6-month follow-up period. | Prospective cohort study  3417 patients aged 10 years and over. | Four of the nine measurements in the Health Literacy Questionnaire (HLQ)  (1) Access  (2) Understanding;  (3) Appraise;  (4) Social | Self-reported measure. | Crude model:  A significant association was found between participants adherence to PA recommendation (≥ 150 mins/wk) and adequate HL: *social support* (Coef. 22.81, 95% CI 15.24-30.39, p < 0.001); *appraisal* (Coef. 14.22, 95% CI 8.03-20.40, p < 0.001); *access* (Coef. 21.73, 95% CI 16.42-27.04, p < 0.001); and *understanding* (Coef. 26.04, 95% CI 19.80-32.28, p < 0.001).  Adjusted*:  A significant association was found between participants adherence to PA recommendation (≥ 150 mins/wk) and adequate HL: *social support* (Coef. 20.55, 95% CI 12.90-28.19, p < 0.001); *appraisal* (Coef. 11.80, 95% CI 5.58-18.02, p < 0.001); *access* (Coef. 17.10, 95% CI 11.35-22.85, p < 0.001); and *understanding* (Coef. 20.51, 95% CI 13.89-27.14, p < 0.001). |
| 5 | Emiral et al.  2021  Turkey | To asses the potential determinates for both knowledge level of metabolic syndrome and health literacy level among the adult population in Western Turkey. | Cross sectional study  774 participants | Health Literacy Scale European Union (HLS-EU-Q16)  (1) Health care  (2) Disease prevention;  (3) Health promotion | Self-reported measure of PA using a single question. | Participants with higher mean HL score (33.40±8.50 vs 31.82±7.02) had a higher prevalence of sufficient PA (≥ 30 min/d). The results were significant (p = 0.011). |

Table 1. (continued)

| **No** | **Authors,**  **Year of Publication,**  **Country** | **Research question** | **Study design**  **Population** | **Health literacy Scale**  **Health literacy**  **domains being measured** | **Measure of PA** | **Results** |
| --- | --- | --- | --- | --- | --- | --- |
| 6 | Goboers et al.  2014  Netherlands | To assess the association of health literacy and physical activity and nutritional behavior in older adults, and to determine if this association is mediated by social cognitive factors. | Longitudinal study  525 community-dwelling older adults (≥ 55 years). | Three validated questions.  (1) Problems learning;  (2) Confidence filling out medical forms;  (3) Help reading. | Self-reported measure of PA using the SQUASH questionnaire | 32.4% of participants with inadequate HL were insufficiently PA (< 150 mins/wk), compared to 21.7% of participants with adequate HL who were insufficiently PA (< 150 mins/wk). The results were significant (p = 0.007).  Crude model:  A significant association was found between insufficiently PA (< 150 mins/wk) and inadequate HL (OR 1.74, 95% CI 1.16-2.95, P = 0.007).  Adjusted model*:  A significant association was found between insufficiently PA (< 150 mins/wk) and inadequate HL (OR_adj_ = 1.52, 95% CI 1.00-2.31, P = 0.020).  Self-efficacy mediated 32% of the association between HL and sufficient PA. Neutral/negative attitude and low risk perception failed mediate association.  * Adjusted for condition, age and gender |
| 7 | Goboers et al.  2016  Netherlands | To assess the associations between health literacy and various health behaviours and social factors among older adults, as well as to investigate whether social factors moderate the associations. | Cohort and case control study.  3241 older adults  (≥ 65 years). | Three validated questions  (1) Problems learning;  (2) Confidence filling out medical forms;  (3) Help reading. | Self-reported measure of PA using a single question. | 33.5% of participants with inadequate HL were insufficiently PA (< 150 mins/wk), compared to 27.6% of participants with adequate HL who were insufficiently PA (< 150 mins/wk). The results were significant (p < 0.01).  Crude:  A significant association was found between insufficiently PA (< 150 mins/wk) and inadequate (OR 1.32, 95% I 1.13-1.55, P < 0.01).  Adjusted*:  A significant association was found between insufficiently PA (< 150 mins/wk) and inadequate HL (OR_adj_ = 1.31, 95% CI 1.11 – 1.54, P < 0.001).  Social factors (Loneliness, Social support, Social activities  Social contacts, and Living situation) failed to mediate association  * Adjusted for age and gender |

Table 1. (continued)

| **No** | **Authors,**  **Year of Publication,**  **Country** | **Research question** | **Study design**  **Population** | **Health literacy Scale**  **Health literacy**  **domains being measured** | **Measure of PA** | **Results** |
| --- | --- | --- | --- | --- | --- | --- |
| 8 | Gibney and Doyle  2017  Ireland | To investigate the relationship between self-rated health literacy and exercise frequency among individuals aged 50+ in Ireland. | Cross sectional study  389 adults aged 50+ | HLS-EU-Q47  (1) Health care  (2) Disease prevention;  (3) Health promotion | Self-reported measure of PA using a single question. | Participants with lower mean HL sub-domain health promotion score (32.0±10.3) had a higher prevalence of insufficient PA (< 150 mins/wk), compared with participants with higher mean HL sub-domain health promotion score (36.2±8.3) had a higher prevalence of sufficient PA (≥ 150 mins/wk). The results were significant (p = 0.010).  Participants with lower mean score for HL sub-domain health promotion: access (6.1.0±2.3) had a higher prevalence of insufficient PA (< 150 mins/wk), compared with participants with higher mean HL sub-domain health promotion: access (7.0±1.8) had a higher prevalence of sufficient PA (≥ 150 mins/wk). The results were significant (p = 0.004).  Adjusted*:  A significant association was found between sufficient PA (≥ 150 mins/wk) and higher HL domains: disease prevention-understanding (OR_adj_ = 1.18, 95% CI 1.03 – 1..34, p < 0.05); health promotion-access (OR_adj_ = 1.13, 95% CI 1.00 – 1..29, p < 0.05); health promotion-understanding (OR_adj_ = 1.15, 95% CI 1.02 – 1..32, p < 0.05); and, health promotion-evaluation (OR_adj_ = 1.13, 95% CI 1.01 – 1..26, p < 0.05).  The non-significant associations and non-associations between sufficient PA (≥ 150 mins/wk) and the other HL indices were not reported.  *Adjusted for age, gender, employment status, marital status, education, financial deprivation, social status and physical limiting illness |

Table 1. (continued)

| **No** | **Authors,**  **Year of Publication,**  **Country** | **Research question** | **Study design**  **Population** | **Health literacy Scale**  **Health literacy**  **domains being measured** | **Measure of PA** | **Results** |
| --- | --- | --- | --- | --- | --- | --- |
| 9 | Jayasinghe et al.  2016  Australia | To investigate the impact of health literacy on Health-Related Quality of Life (HRQoL) | Cross sectional study  379 participants | Health Literacy Management Scale (HeLMS)  (1) Attitudes;  (2) Understand;  (3) Social support;  (4) Socioeconomic  considerations;  (5) Assess;  (6) Interactive;  (7) Being proactive;  (8) Apply. | Self-reported measures | 62.7% of participants with inadequate HL were insufficiently PA (< 150 mins/wk), compared to 48% participants with adequate HL who were insufficiently PA (< 150 mins/wk). The results were significant (p < 0.001).  . |
| 10 | Jennings et al.  2013  Multi-national (Europe) | To explore the association between illness perceptions and health literacy with sociodemographic characteristics and risk factors, health-related quality of life, anxiety and depression. | Cross sectional study  3408 participants (aged 18-80 years) with heart conditions. | Only the 9^th^ scale of the Health Literacy Questionnaire (HLQ-9) was used.  (1) Understanding health information. | Self-reported measures | Participants with lower HL levels were significantly associated with insufficient PA (p < 0.001). |
| 11 | Joshi et al.  2014  Australia | To compare primary car patients with and without sufficient health literacy in terms of their lifestyle risk factors and explore associated with receiving advice and referral for the risk factors from their GP. | Cross sectional study  739 participants | HeLMS  (1) Attitudes;  (2) Understand;  (3) Social support;  (4) Socioeconomic  considerations;  (5) Assess;  (6) Interactive;  (7) Being proactive; | Self-reported measures | A significant association was found between insufficient PA (< 150 mins/wk) and participants with inadequate HL (OR_adj_ = 1.81, 95% CI 1.34 – 2.43, p < 0.001). |
| 12 | Jürgensen et al.  2024  Germany | To explore and describe the health status, health-related behavior, and health literacy levels of first-year health professional students, as well as to investigate potential differences between two groups of students. | Cross sectional study  90 first year health professional students | HLS-EU-Q16  (1) Health care  (2) Disease prevention;  (3) Health promotion  . | Self-reported measures | 65.1% of participants with inadequate HL were insufficiently PA (< 2h/wk), compared to 61.5% participants with adequate HL who were insufficiently PA (< 2h/wk). The results were not significant (p = 0.831). |

Table 1. (continued)

| **No** | **Authors,**  **Year of Publication,**  **Country** | **Research question** | **Study design**  **Population** | **Health literacy Scale**  **Health literacy**  **domains being measured** | **Measure of PA** | **Results** |
| --- | --- | --- | --- | --- | --- | --- |
| 13 | Juul et al.  2018  Denmark | To investigate associations between health literacy and diet and physical activity, and motivation and diet and physical activity in Danish people with type 2 diabetes. | Cross sectional study  194 adults with type 2 diabetes | HLS-EU-Q16  (1) Functional;  (2) Critical;  (3) Communicative | Self-reported measures using the SDSCA. | Crude:  A non-significant association was found between frequency of achieving sufficient PA (≥ 30min/d) and functional HL (β 0.31, 95% CI -0.12-0.74), and communicative HL (β 0.38, 95% CI -0.16-0.92).  No association was found between frequency of achieving sufficient PA (≥ 30min/d) and total HL (β 0.06, 95% CI -0.12-0.74), and critical HL (≥ 30min/d) (β 0.17, 95% CI -0.33-0.67).  Adjusted:  No association was found between total HL, functional HL, communicative HL or critical HL and frequency of achieving sufficient PA (≥ 30min/d).  *Adjusted for age, gender education level, diabetes duration, and other HL scores. |
| 14 | Kobayashi et al.  2016  United Kingdom | To investigate the relationship between health literacy and participation in weekly moderate to vigorous physical activity over an 8-year period among older adults. | Longitudinal study  4345 adults aged 52-79 from the English Longitudinal Study | Validated four-item measure from the Organisation For Economic Co-operation and Development Internation Adult Literacy Survey  (1) Reading comprehension | Self-reported measures | Crude:  An association was found between sufficient PA (≥ 150 mins/wk)and participants with medium HL (OR 1.83, 95% CI 1.41-2.37); and high HL (OR 2.83, 95% CI 2.25-3.57)  Adjusted*:  An association was found sufficient PA (≥ 150 mins/wk) and participants with medium HL (OR 1.29, 95% CI 0.95-1.75), and high HL (< 150 mins/wk) (OR 1.53, 95% CI 1.16-2.01).  Adjusted**:  An association was found between sufficient PA (≥ 150 mins/wk) and participants with medium HL (OR 1.21, 95% CI 0.89-1.64), and high HL (OR 1.37, 95% CI 1.04-1.81).  *Adjusted for age, sex, education attainment, net non-person wealth, self-rated health, limiting long-standing illness, and IADL limitation compliance  **Additional adjustments for memory an verbal fluency. |

Table 1. (continued)

| **No** | **Authors,**  **Year of Publication,**  **Country** | **Research question** | **Study design**  **Population** | **Health literacy Scale**  **Health literacy**  **domains being measured** | **Measure of PA** | **Results** |
| --- | --- | --- | --- | --- | --- | --- |
| 15 | Koch et al.  2023  Germany | To monitor the health of trainees in different sectors over time, ending during the COVID-19 pandemic, and to examine the association between health literacy and health/health behaviour. | Longitudinal study  129 trainees from various sectors. | HLS-EU-Q16  (1) Health care  (2) Disease prevention;  (3) Health promotion | Self-reported measures | No association was found between HL (at baseline) and PA (3 months) (p = 0.124). |
| 16 | Nagaki et al.  2023  Japan | To evaluate the association between physical activity levels and different health literacy domains in patients with Parkinsons disease. | Cross sectional study  141 Japanese patients with Parkinsons disease aged 18 years or older. | Functional, Communicative, ad Critical Health Literacy (FCCHL) scale.  (1) Functional;  (2) Critical;  (3) Communicative | Self-reported measures using the IPAQ Short Form. | Crude model:  A significant association was found between participants adherence to PA recommendation (≥ 150 mins/wk) and Critical HL (OR 2.12, 95% CI 1.06-4.22, p = 0.03).  A non-significant association was found between participants adherence to PA recommendation (≥ 150 mins/wk) and functional HL (OR 1.40, 95% CI 0.77-2.53, p = 0.26) and communicative HL (OR 1.42, 95% CI 0.65-3.11, p = 0.38).  Adjudged model^*^:  A significant association was found between participants adherence to PA recommendation (≥ 150 mins/wk) and Critical HL (OR_adj_ 2.46, 95% CI 1.16-5.19, p = 0.02).  A non-significant association was found between participants adherence to PA recommendation (≥ 150 mins/wk) and functional HL (OR_adj_ 1.58, 95% CI 0.84-2.96, p = 0.16). and communicative HL (OR_adj_ 1.51, 95% CI 0.66-5.19, p = 0.38).  * Adjusted for age, gender, PD duration, and academic background. |
| 17 | Reider et al  2017  United States of America | To determine whether health literacy was associated with meeting the recommended guidelines for physical activity in people with multiple sclerosis. | Cross sectional study  9019 adults with Multiple Sclerosis | METER  (1) Functional health literacy | Self-reported measures using the Health Information National Trends (HINTS) survey | No association was found between adherence to PA recommendation (≥ 150 mins/wk) and Functional HL (OR 0.91, 95% CI 0.78-1.06, p > 0.05). |

Table 1. (continued)

| **No** | **Authors,**  **Year of Publication,**  **Country** | **Research question** | **Study design**  **Population** | **Health literacy Scale**  **Health literacy**  **domains being measured** | **Measure of PA** | **Results** |
| --- | --- | --- | --- | --- | --- | --- |
| 18 | Shahavandi et al  2021  Iran | To evaluate the associations between health literacy and healthy eating index (HEI) in adults. | Cross-sectional study  261 Iranian adults aged 18 to 65 years. | Health Literacy for Iranian Adults (HELIA) questionnaire.  (1) Accessing;  (2) Reading;  (3) Comprehension;  (4) Assessment;  (5) Decision-making | Self-reported measures using the IPAQ Short Form. | A significant association was found between adherence to PA recommendation (≥ 150 mins/wk) and HL (β 0.12, 95% CI -0.05-1.72, p = 0.03). |
| 29 | Zanobini et al  2021  Italy | To evaluate health literacy levels and their associations with sociodemographic factors and health-related behaviours in a representative sample of the Tuscan population | Cross-sectional study  7151 adults | The Italian version of the HLS-EU-Q6  (1) Health care  (2) Disease prevention;  (3) Health promotion | Self-reported measures | A significant association was found between adherence to PA recommendation (≥ 150 mins/wk) and HL (p <0.001).  Adjudged model^*^:  A significant association was found between participants adherence to PA recommendation (≥ 150 mins/wk) and HL (OR_adj_ 0.60, 95% CI 0.50–0.73, p = 0.000).  *Adjusted for age, sex, educational level, occupation status,  financial status and nationality |
